# Study of the recommended dosage of the N-Acetyl Cysteine, Alpha Lipoic Acid, Bromelain and Zinc preparation as a treatment for dysmenorrhea

**DOI:** 10.1101/2022.08.08.22278399

**Authors:** Zuramis Estrada, Francisco Carmona

**Author notes:** **Contact information:** Francisco Carmona, MD, PhD, Endometriosis Unit, ICGON, Hospital Clinic of Barcelona, University of Barcelona, Villarroel 170, 08036 Barcelona, Spain.

## Abstract

1.

**AIM:** Dysmenorrhea, defined as pain during menstruation, is the most common gynecological condition, affecting a large percentage of women with varying degrees of pain. In recent years, the management of dysmenorrhea has become increasingly important because of the emotional, health and economic burden it entails and because of the need for new studies and research for effective treatments to alleviate its symptoms. The most common treatments for dysmenorrhea are NSAIDs or oral contraceptives, although gynecologists also recommend the preparation composed of N-acetyl cysteine (NAC), alpha lipoic acid (LA), bromelain (Br) and Zinc (Zn), (NAC/LA/Br/Zn) due to its anti-inflammatory and anti-oxidative properties and efficacy in reducing pain. However, there is no data on what dosage of the preparation should be recommended for patients with dysmenorrhea. Therefore, the aim of this study is to determine how the NAC/LA/Br/Zn preparation is recommended for the treatment of dysmenorrhea in daily clinical practice and what specialists base their decision on.

**Methods and Results:** A survey was conducted among gynecologists with extensive experience recommending the preparation, with a participation rate of 97% (N=73). The most frequently recommended regimen is to begin with continuous administration for 90 days with 10-day breaks (69%) or without breaks (31%). Specialists recommend the preparation for any type of pain and may recommend discontinuous administration (in the days around menstruation) when the pain is moderate or mild. In patients with primary dysmenorrhea with mild pain, the most recommended approach is discontinuous therapy in the days around menstruation (50.7%) followed by continuous administration for 90 days with 10-day breaks (24.7%). When pain is moderate or severe, the most recommended approach is 90-day continuous therapy with 10-day breaks (47.9% and 71.2%, respectively). In the case of secondary dysmenorrhea, the most recommended approach is 90-day continuous therapy with 10-day breaks for any degree of pain (41.1%, 57.5% and 76.7% for mild, moderate and severe pain, respectively). Most gynecologists (79%) adapt the regimen after clinical assessment of the degree of pain towards discontinuous administration patterns, on the days around menstruation. Most of the specialists consulted do not modify the recommended regimen if the patient is being treated with other drugs such as NSAIDs, hormonal contraception or a combination of progestogens (78%, 59% and 58%, respectively). The preparation is recommended in conjunction with hygienic-dietary measures without modification of the dosage (90%).

**Conclusion:** This is the first study that addresses how specialists recommend the NAC/LA/Br/Zn preparation to patients with dysmenorrhea. The most common regimen for dysmenorrhea is to start treatment with continuous administration of the preparation for 90 days with 10 days of break, continuing the regimen or adapting it according to the degree of pain reported by the patient during treatment.

## 2. Introduction

Dysmenorrhea, defined as pain during menstruation (1,2), is the most common gynecological condition, affecting a large percentage of women with varying degrees of pain (3,4). Although there is no consensus in the literature on the prevalence of dysmenorrhea, its prevalence lies between 17 and 81% of women, with severe dysmenorrhea occurring in 12 to 14% (5). Dysmenorrhea is classified as primary, when the underlying anomaly is unknown, or secondary, which is usually strongly associated with endometriosis or pelvic inflammatory disease, among other causes (4,6-16). In recent years, the management of dysmenorrhea has become increasingly important because of the emotional, health and economic burden it entails and because of the need for new studies and research for effective treatments to alleviate its symptoms (1,5,17).

Treatments for dysmenorrhea can be classified as pharmacological, such as non-steroidal anti-inflammatory drugs (NSAIDs) and combined hormonal contraceptives (18-22), or non-pharmacological, such as complementary treatments to reduce pain (23-26). Currently, gynecologists recommend the preparation of N-acetyl cysteine (NAC), alpha lipoic acid (LA), bromelain (Br) and zinc (Zn) (NAC/LA/Br/Zn), due to its antioxidant, anti-inflammatory and analgesic properties (27,28).

Due to the lack of evidence on how the NAC/LA/Br/Zn preparation is used in daily clinical practice, we decided to conduct a survey to gather information on the experience of gynecologists who recommend it as a treatment for dysmenorrhea. Therefore, the aim of the study is to find out the most frequent doses of the preparation recommended for both primary dysmenorrhea (PD) and secondary dysmenorrhea (SD) and other causes of pain in relation to the reasons for recommending it, the characteristics of the patients and the adaptation according to the response to treatment.

## 3. Materials and Methods

### 3.1. Survey approach

The study involved a total of 75 gynecologists with extensive professional experience in the treatment of dysmenorrhea, from different geographical areas of Spain. The survey developed was sent to gynecologists with experience treating patients with the preparation of N-acetyl cysteine (NAC), alpha lipoic acid (LA), bromelain (Br), Zinc (Zn). All identities were kept confidential during data collection and analysis. The participation of the specialists was voluntary, free and unconditional.

The data on treatment times and concentration of preparation components reported in patients with endometriosis by Lete *et al*. (27) were used as a reference for developing the survey. The specialists were invited to participate by responding to a structured survey aimed at revealing the recommended dosage of the NAC/LA/Br/Zn preparation for patients with menstrual pain. The survey was structured in five sections: [1] previous considerations (reason for use of the preparation and patient characteristics) [2] Primary Dysmenorrhea (patient management and dosage) [3] Secondary Dysmenorrhea and other causes of pain (patient management and dosage); [4] association of the NAC/LA/Br/Zn preparation with other drugs; [5] final considerations and comments from the specialist consulted. The survey consisted of 37 questions in total, 32 of which were single-answer questions, 5 of which were multiple-choice questions. In all questions, open text fields were available to enter responses not included in the list, except in five questions where the proposed options were categorical. The complete survey can be found in the supplementary material.

### 3.2. Data analysis and statistics

Gynecologists’ responses were collected using the Google Forms tool (Google LLC), and downloaded into a Microsoft Excel 2019 file (Microsoft Office, Redmond, WA) for analysis. Open-ended questions were manually coded into general response categories by a single author using inductive coding with iterative sampling and recoding. In the multiple-choice questions, the number of times each answer was selected was analyzed. None of the participants’ responses were excluded from the analysis. Descriptive statistics including absolute numbers, frequencies (%) and means were used to explore the data.

## 4. Results

A high level of participation was attained from the gynecologists identified, 73 responses out of the 75 invited to participate (97.3%) having completed all sections of the survey. The gynecologists consulted recommend the NAC/LA/Br/Zn preparation to patients with any cause of menstrual pain, especially to women who want to reduce the consumption of NSAIDs (48 responses), who wish to become pregnant (46 responses), and patients with pain who do not want to take contraceptives (32 responses) (possibility of multiple responses) (figure 1A). The majority of gynecologists recommend the NAC/LA/Br/Zn preparation to patients who report any degree of pain (49%), followed by the recommendation to patients who report moderate pain (between 4 and 7 pain on the *Visual Analogue Scale* (VAS), 37%) or patients with severe pain (>7 VAS, 14%) (Figure 1B).

**Figure 1.**
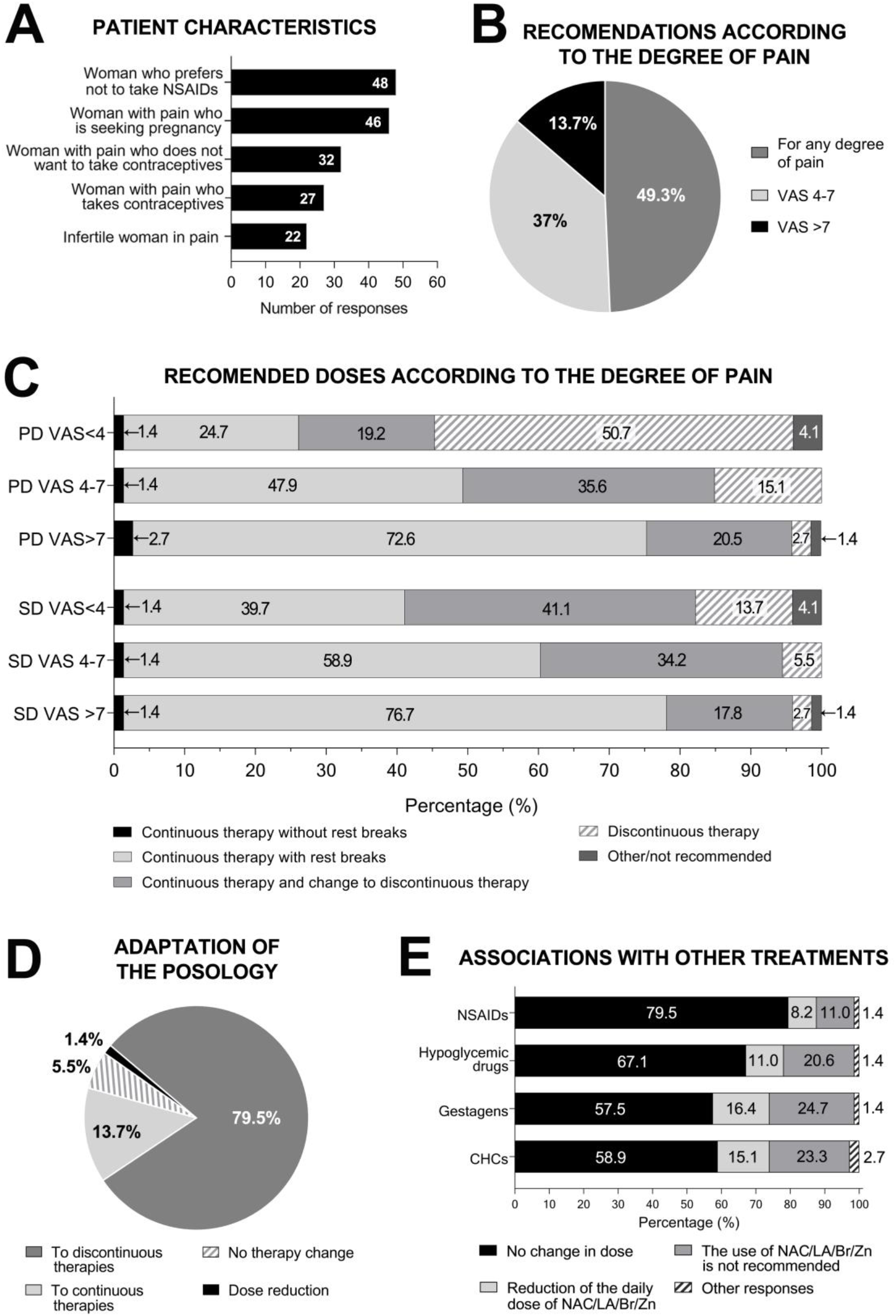
Recommended dosage of NAC/LA/Br/Zn with respect to patient characteristics, degree of pain and associations with other treatments. **(A)** Expert opinion on the indication for which the preparation is recommended regarding the patient characteristics, **(B)** recommendations of the preparation according to pain reported by the patient following the VAS scale (Visual Analogue Scale, from 0 to 10). **(C)** Recommended posology according to the disease diagnosed (PD: upper bars; SD: bottom bars) and the pain reported by the patients following the VAS scale. **(D)** Adaptation of the posology in PD according to expert opinion and the patient report to discontinued therapies 79%; to continuous therapies 14%; no therapy changes 5%; dose reduction 1%). **(E)** Associations with treatments (NSAIDs: Non-steroidal anti-inflammatory drugs; CHCs: Combined hormonal contraception). (A) was posed as multiple-choice questions; the number of votes for each answer is shown. (B, C, D and E) responses are represented as a percentage of responses, N=73. Percentages are shown in the figures. PD: primary dysmenorrhea. SD: secondary dysmenorrhea.

### 4.1. Recommended Dosages

At the beginning of treatment with the preparation, gynecologists recommend the administration of NAC 600 mg/day, LA 200 mg/day, Br 25 mg/day Zn 10 mg/day (71.2% for PD, 80.8% for SD), while the rest start treatment with half the concentration, with the possibility of doubling the dose if the patient does not respond to treatment (27.8% for PD and 19.2% for SD) (Figure 1A supplementary material).

The regimens recommended by specialists at the beginning of treatment may be continuous or discontinuous. The most commonly recommended continuous regimen is continuous therapy with breaks (10 days of rest per 90-day treatment cycle) recommended by 69% of the specialists, compared to continuous therapy without breaks (31% of the specialists) (Table 1). Discontinuous regimens entail administration of the preparation 7-15 days before menstruation (54%), 5 days before and 5 days after menstruation (24%), only 5 days before menstruation (15%) or other variations (7%) (Table 1).

**Table 1:**
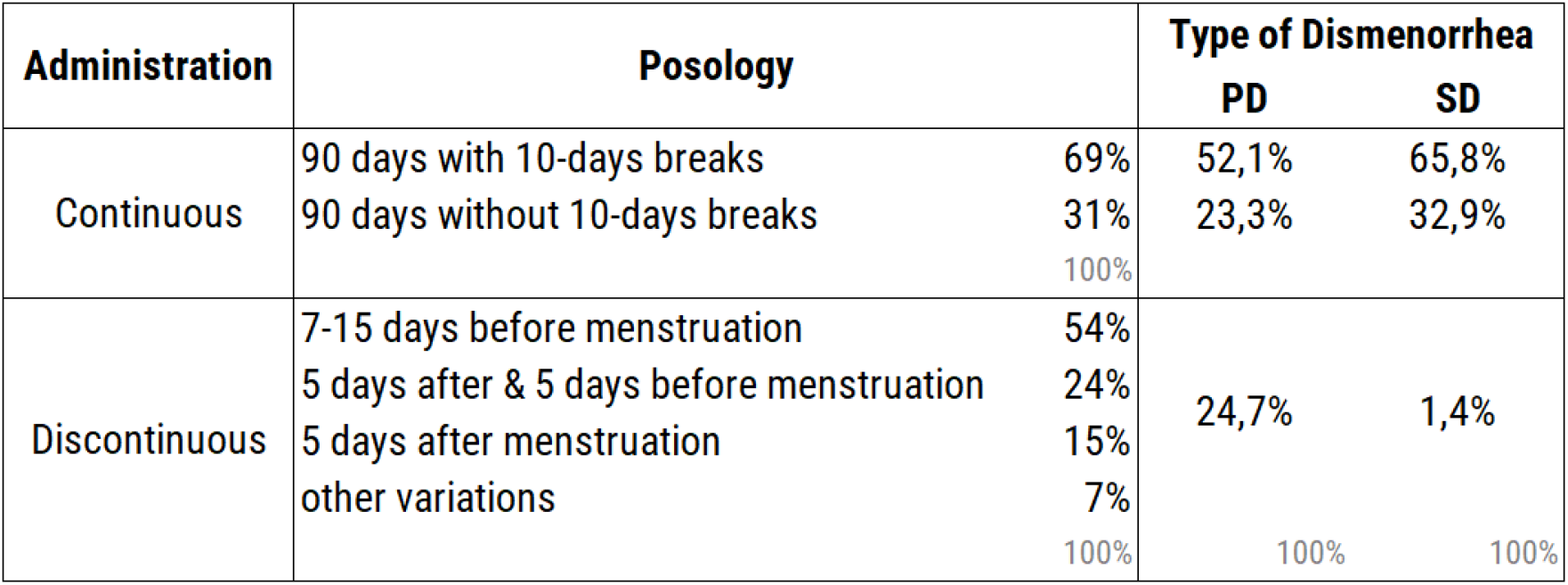
Recommended dosages of the NAC/LA/Br/Zn preparation at the start of treatment of dysmenorrhea. N=73.

For the treatment of dysmenorrhea, the most commonly recommended therapy at the start of treatment is continuous administration with 10-day breaks every 90 days of treatment (54.8% for PD and 65.8% for SD). In the case of PD, 34.2% recommend long-duration therapies (>90 days) and 11% short-duration (<90 days). In the case of SD, 32.9% also recommend continuous administration without breaks at the start of treatment, and 1.4% recommend short-duration therapies (<90 days).

Gynecologists decide the regimen they recommend based on the pain reported by patients. For PD patients with mild pain, most specialists recommend discontinuous therapies (50.7%), followed by continuous therapy with breaks (24.7%) and administration for 90 days with change to discontinuous therapy (19.2%). For patients with moderate or severe pain, the most recommended option is continuous therapy with breaks (47.9% and 72.6%, respectively), followed by administration for 90 days with change to discontinuous (35.6% and 20.5%, respectively) (Figure 1C and Table 2).

**Table 2.**
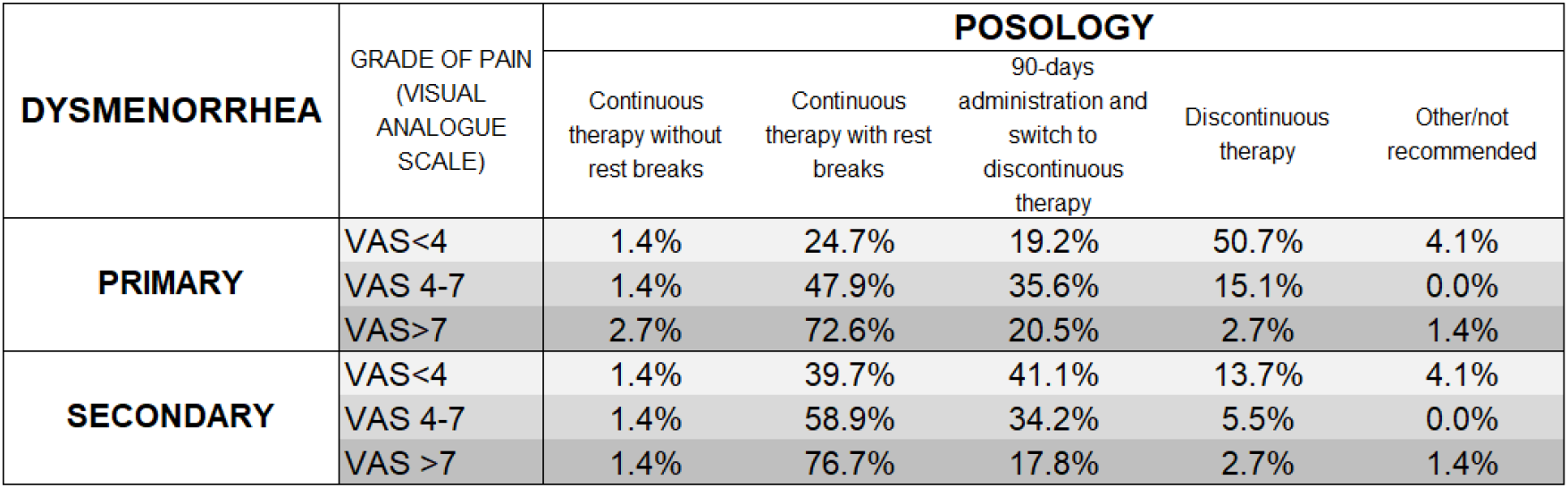
Recommended posology according to the disease diagnosed and the pain reported by the patients following the VAS scale. [1] Continuous therapy without breaks; [2] Continuous therapy with breaks (10 days off per 90-day treatment cycle); [3] 90-days administration and switch to discontinuous therapy; [4] Discontinuous therapy (5 days before menstruation and 5 days after or 7-15 days before menstruation). N=73.

In contrast to PD, for SD patients, for patients with mild pain, most specialist recommended 90 days with change to discontinuous (41,1%), followed by continuous therapy with breaks (39.7%) and discontinuous therapy (13.7%) (Figure 1C and Table 2). For patients with moderate or severe pain, the most recommended option is continuous therapy with breaks (58.9% and 76.7%, respectively), followed by administration for 90 days with change to discontinuous (34.2% and 17.8%, respectively) (Figure 1C and Table 2).

For other causes of pain, 63% of gynecologists recommend the preparation, while 33% only recommend it for endometriosis or dysmenorrhea associated with endometriosis, and the remaining 4% recommend it when all other treatments fail. Among those who do recommend the preparation, the most commonly recommended dosage is continuous therapy with 10-day breaks (47.4%), followed by administration for 90 days with a change to discontinuous therapy (31.6%), and 17.6% recommend discontinuous therapies (7-15 days before menstruation or 5 days before and 5 days after, both recommended with the same frequency).

Regarding dosage adaptation, 79.5% of gynecologists recommend switching from continuous to discontinuous administration, while other gynecologists recommend switching from discontinuous to continuous administration (13.7%), not adapting the regimen (5.5%) or choosing other options (1.4%) (Figure 1D). Among specialists who recommend switching to discontinued therapies, the most recommended regimens are administration in the 7-15 days before menstruation (44%), 5 days before (19%) or 5 days before and after menstruation (16%).

### 4.2. Association of the preparation with other pharmaceuticals

Some patients who are recommended the NAC/LA/Br/Zn preparation are being treated with other pharmaceuticals. Gynecologists recommend the preparation without modification of the dosage when they are treated with NSAIDs, gestagens or the combination of hormonal contraception (79.5%, 57.5% and 58.9%, respectively), compared to those who do modify the dosage of the preparation (8.2%, 16.4% and 15.1%, respectively). Some specialists do not recommend the preparation when the patient is being treated with NSAIDs, gestagens, combination of hormonal contraceptives (11%, 24.7% and 23.3% respectively). In cases in which patients are being treated with hypoglycemic drugs, 67.1% of gynecologists recommend the NAC/LA/Br/Zn preparation without modifying the regimen, compared to 20.6% who do not recommend it in these cases and 11% who reduce the daily dose (Figure 1E supplementary material).

### 4.3. Combined use with hygienic-sanitary measures

In most cases, in addition to treatment, patients are recommended a series of hygienic-dietary measures, based on healthy eating, exercise and avoiding stress. Ninety percent of gynecologists recommend the NAC/LA/Br/Zn preparation along with all hygienic-dietary measures, 6% recommend the preparation alone and 4% recommend the preparation with a healthy diet. Ninety percent of specialists do not modify the dosage of the preparation when recommending hygienic-dietary guidelines (1C supplementary material).

### 4.4. Patient satisfaction and reported adverse effects

Depending on the progression of pain reported by patients, 92% adapt the dosage of the preparation compared to those who do not (8%), adaptation to discontinuous treatment being the most frequent. For gynecologists who do adapt the dosage, the most frequent change is from continuous to discontinuous treatments (60%) compared to those who report changes from discontinuous to continuous regimens (26%). Finally, the participants in this study stated that the NAC/LA/Br/Zn preparation is well tolerated by patients, with gastrointestinal discomfort being the only adverse effect, detected by 11% of the gynecologists consulted (Figure 1D supplementary material).

## 5. Discussion

This study describes for the first time which dosages of the NAC/LA/Br/Zn preparation are most recommended by gynecological experts for patients with dysmenorrhea. The most frequently recommended regimen is to begin with continuous administration for 90 days with 10-day breaks. In subsequent evaluations, specialists may maintain this dosage or change it if the patient reports a reduction in pain. In addition, this study sheds light on how gynecologists recommend the NAC/LA/Br/Zn preparation depending on patients’ characteristics, degree of pain, the type of dysmenorrhea and the association with other pharmaceuticals.

Given the lack of data in the literature on the guidelines for recommending the preparation in clinical practice, it is necessary to conduct studies to produce pertinent data. It is important to know the opinion of gynecologists who have experience recommending the preparation to patients with dysmenorrhea in order to learn how they prescribe it. The chosen methodology, an approach validated and used in other clinical areas, was a survey of gynecologists in Spain (29– 31).

At the beginning of the treatment for dysmenorrhea, the most frequently recommended regimen is continuous administration for 90 days with 10-day breaks, although one third of those consulted recommend continuous administration without breaks. In subsequent evaluations, the specialist may recommend continuing with this regimen or changing it to a discontinuous dosage if the pain has subsided. In the instance of patients who initially report mild pain, the specialist may recommend starting therapy with a discontinuous administration of the preparation. The most frequently recommended discontinuous regimens involve administering the preparation 7-15 days before menstruation or 5 days before and 5 days after. Administering the preparation around the days of menstruation results in a reduction in menstrual pain. Some specialists have stated that it is possible to switch to a continuous dosage in the event that the patient reports an increase in pain following a discontinuous dosage.

For primary dysmenorrhea, gynecologists recommend and adapt the dosage of the preparation according to the patient’s pain. When the pain is moderate or severe, gynecologists mostly opt for continuous treatment, and when the pain is mild, discontinuous treatment is more frequently recommended. Upon reevaluation, the tendency is to reduce the number of doses of the preparation because it continues to be effective in reducing pain, although in some cases it can be changed from a discontinuous to a continuous regimen if the patient reports increased pain with a discontinuous regimen. However, in secondary dysmenorrhea, for patients with any degree of pain, the most recommended guideline is continuous therapy followed by re-evaluation. This may be due to the fact that in secondary dysmenorrhea the patient has been previously diagnosed with endometriosis, which is the probable cause of the pain, so gynecologists more often recommend long-acting regimens to reduce pain following the dosage reported in the study by Lete *et al* (27). This study also addressed the issue of how the preparation is recommended for patients with other causes of pelvic pain, since it was also found to be effective in reducing their pain. The most commonly recommended therapy for other causes of pelvic pain, as well as for PD and SD, is the administration of the preparation for 90 days with 10-day breaks.

There is evidence of the effectiveness of this preparation in patients diagnosed with pelvic pain associated with endometriosis when the regimen is NAC 600 mg/day, LA 200 mg/day, Br 25 mg/day and Zn 10 mg/day for 6 months (27). In this prospective, open-label clinical study, it was observed that patients taking the preparation reported a reduction in pain from 6.7 to 3.5 on the VAS scale and a reduction in the intake of NSAIDs, whether regularly administered or on menstrual days. The evidence available to date indicates that the individual compounds separately are beneficial for patients with endometriosis-associated menstrual pain, but at different doses or treatment times than those recommended in daily clinical practice. Treatment with NAC significantly reduced pain caused by dysmenorrhea and vaginal bleeding when administered at the same concentration as in this study for 90 days (32,33). Lower doses of NAC (150 mg/day), short treatments (60 days) and in combination with other molecules also favor the reduction of painful symptoms associated with endometriosis (34). LA as a treatment for dysmenorrhea and chronic pelvic pain associated with endometriosis reported greater efficacy in reducing pain when administered in combination with other compounds (35). Continuous treatment and treatment on the days before menstruation have shown positive results in reducing pain (35,36). A *case report* describes that administration of Br (2000 mg/day changing to 3000 mg/day 3 days before menses) in combination with other compounds and acupuncture significantly reduced pain (from 8 to 2 on the VAS scale) (37). Other studies administering bromelain extract (1.5 mg/mL in PBS, 5 minutes vaginal irrigation) showed immediate relief in 62% of patients (38,39). Zn supplementation has been reported to prevent uterine spasms and cramps, inducing a reduction in prostaglandin synthesis by reducing cyclooxygenase-2 (COX-2) activity and an improvement in endometrial tissue microcirculation (40-42). The administration of Zinc, at concentrations between 20 and 126 mg/day for 3-6 days before and after menstruation, reduces pain in the first menstrual cycle after the first intake, although the reduction in pain was greater in subsequent cycles (42–45). Regarding dosage, concentrations higher than 30 mg/day did not report better additional effect, probably due to the poor absorption of zinc in the intestine (42,46).

Numerous therapeutic strategies and drugs approved by regulatory agencies are currently available to treat endometriosis and associated symptoms such as dysmenorrhea. In most cases, specialists recommend the preparation in conjunction with the prescribed treatment (between 60% and 80%) as opposed to those who do not (between 10% and 25%). Gynecologists more frequently recommend the preparation when it is associated with NSAIDs, probably because of the anti-inflammatory properties of the preparation and because it can complement or reduce the administration of NSAIDs (27). Less frequently, they recommend the preparation when the patient is being treated with hormonal contraception or with a combination of progestogens. This may be because oral contraceptive treatments are able to inhibit ovulation and endometrial proliferation, reducing or eliminating menstrual pain, resulting in specialists recommending the combination less frequently in these cases.

Clinical guidelines for the management of patients with menstrual pain also include some non-pharmacological strategies or recommendations, such as exercise, healthy diet and natural remedies. Although there is no solid scientific evidence in this regard, the components of the preparation have antioxidant, anti-inflammatory and analgesic properties (35,41,47). Therefore, specialists recommend the preparation for its effectiveness in reducing pain as a complement to promoting healthy habits.

The NAC/LA/Br/Zn preparation is well tolerated by patients, with adverse reactions reported in only 11% of patients. Adverse reactions reported were mild gastrointestinal discomfort, such as epigastralgia, gastralgia, abdominal bloating, gastric intolerance or heartburn. Among the components of the preparation, gastric problems such as nausea, vomiting, diarrhea, pain and heartburn have been reported as adverse effects of NAC, occurring infrequently (1:1000) (48).

## 6. Conclusions

The most common regimen for the treatment of dysmenorrhea with the NAC/LA/Br/Zn preparation (600 mg/day, 200 mg/day, 25 mg/day and 10 mg/day) is to start treatment with 90 days of administration with 10-day breaks. In subsequent evaluations with the specialist, the treatment can be continued with the same continuous regimen with breaks or adapted according to the degree of pain. In the case of adapting the pattern to a discontinuous dosage, the regimen most frequently recommended by specialists is the administration of the preparation 7-15 days before menstruation.

The study obtained a wide participation of gynecologists who are experienced in treating dysmenorrhea and who recommend the NAC/LA/Br/Zn preparation in daily clinical practice. The gynecologists responded freely and without any prior consideration, including all their answers in the analysis. A lot of information has been collected on how the preparation is recommended for patients with any degree of pain at the beginning of treatment and after re-evaluations, as well as the association with other treatments or therapies. In addition, details were obtained on how patients with dysmenorrhea are evaluated, segmented and followed up so that they receive the most appropriate dosage. The results obtained would justify and support further studies on the effectiveness of the preparation in patients with dysmenorrhea.

### Limitations

This study did not include the testimony of patients, nor clinical data of patients with dysmenorrhea who are being treated with the preparation. Due to the main objective of the study, the survey was directed specially to gynecologists who had experience recommending the preparation.

## Supporting information

Survey

## Data Availability

All data produced in the present study are available upon reasonable request to the authors

## Conflicts of interest

The authors declare they have no conflict of interest.

## Acknowledgements

The study was funded by a grant from Adamed Spain S.L.U, who was not involved in the design of the study, and data collection or interpretation.

The authors would like to thank our colleagues that responded to the survey: I. Adiego Calvo; M. Al Adib Mendiri; R. Alania López; A. Alonso García; C. Alonso García; J. Armengol Santacreu; A. Bahillo Varela; R. Baltà i Arandes; R. M. Barceló Tortella; A. I. Barqueros Ramírez; R. Barrientos Naz; A. Biterna Tejeiro; I. Bonal Cea; P. Burguete Fenollosa; E. S. Cabo Silva; R. Campos Caballero; A. Cañadas Molina; A. Carazo Piñeiro; A. Carballo García; M. J. Carballo Martínez; M. Carrascoso Altares; T. Casanova Sanchis; A. Cearsolo Michelena; C. Chacón Aguilar; A. Chassignet Martin; M. C. Chicharro Cassuso; R. Curiel Rodado; J. Dapena González; Miriam de la Flor López; Lucia Diaz Meca; V. Domínguez Rubio; M. J. Fernández Ramírez; E. Fresnadillo Humet; R. Galvan García; O. Gómez Pardo; N. Fernández Aller; M. González Jareño; E. González Rodríguez; C. Grau Bravo; E. Grau Civit; F. Hamadache Chabouni; R. Herrera Recio; A. Jodar Santibáñez; M. Juárez Cuervo; I. Juárez Pallarés; S. Landeo Agüero; M. Lapresta Moros; E. López Pérez; J. López Pérez; D. Lubian López; C. Marcos Santos; C. Miyares Erausquin; F. Moreno Aguayo; M. J. Moreno Pérez; T. Muñoz Fernández; I. Navarro Alonso; M. C. Paladino Decile; M. I. Parra Ribes; M. D. Pérez -Jaraíz López-Zaballa; A. Prades Sanchis; C. Puertas; M. A. Quiñones Chávez; B. Ramos Balbona; R. Reboredo García; E. Recari Elizalde; F. Rubio Fernández; E. Ruipérez Pacheco; J. Sánchez Orta; C. Caninzzo Naccarato; J.F. Subiris González; C. Troncoso Miranda.

## Notes

### Competing Interest Statement

The authors have declared no competing interest.

### Funding Statement

This study was funded by Adamed Spain S.L.U, who was not involved in the design of the study, and data collection or interpretation. The funding includes medical writing of the manuscript and the article processing charges.

