## Supplementary material for "Study of the recommended dosage of the N-Acetyl Cysteine, Alpha Lipoic Acid, Bromelain and Zinc preparation as a treatment for dysmenorrhea": Survey

**PURPOSE OF THE SURVEY**

The main objective of the following survey is to know the recommended posology of the prepared composed by N-acetyl cysteine (NAC), alpha lipoic acid (LA), bromelain (Br) and Zinc (Zn), (NAC/LA/Br/Zn) in patients presenting dysmenorrhea. NAC/LA/Br/Zn is indicated to relieve chronic pelvic pain associated with endometriosis, with a triple action: antioxidant, anti-inflammatory and analgesic.

Dysmenorrhea is defined as uterine pain that takes place at the time of menstruation. It can occur during menstruations or 1 to 3 days before it begins. The pain tends to be more intense 24 hours after the onset of menstruation and continues for 2 to 3 days. Primary dysmenorrhea is defined as pain during menstruation in the absence of an identifiable origin, while secondary dysmenorrhea is often associated with endometriosis or pelvic anatomical abnormalities.

The survey aims to review the possible treatment guidelines depending on the characteristics of the patients, the degree of pain they experience, as well as the adaptation of the posology during the treatment depend on the evolution of the patients. The secondary objective is to know the recommended dosage of the product regarding the type of dysmenorrhea (primary or secondary) and other causes of pain, together with the evaluation of the patient’s satisfaction with the recommended therapeutic regimen.

**SURVEY**

The survey consists of 38 questions divided into 5 blocks of questions, that could be multiple-choice or single-choice answer. In many questions you can write your own answer if you do not agree with the options given. The estimated time required to complete the questionnaire is less than 15 minutes.

**BLOCK 1: PRELIMINARY CONSIDERATIONS**

1. For which **indication** do you recommend the prepared NAC/LA/Br/Zn most frequently? (multiple-choice)
   a. Primary dysmenorrhea
   b. Secondary dysmenorrhea
   c. Other causes of pelvic pain
   d. Other_____________________
2. What is the **main reason** for recommending the prepared NAC/LA/Br/Zn? (multiple-choice)
a. Antioxidant
b. Anti-inflammatory
c. Analgesic
d. Contributes to DNA synthesis
e. The combination of all of them
3. What are the **general characteristics** **of the patients** to whom you recommend the prepared NAC/LA/Br/Zn? (multiple-choice)
   a. Woman in pain who doesn't want to take birth control pills
   b. Woman in pain taking birth control pills
   c. Woman in pain who is looking for a pregnancy
   d. Sterile woman with pain
   e. Woman who prefers not to take NSAIDs/analgesics
   f. Other_____________________

**BLOCK 2: PRIMARY DYSMENORRHEA**

4. Before recommending the prepared NAC/LA/Br/Zn, do you **assess the type** of dysmenorrhea? (Single-choice)
   a. Yes, always
   b. Whenever I can
   c. No
   d. Other_____________________
5. How do you **evaluate** primary dysmenorrhea? (multiple-choice)
   a. Interrogation or anamnesis
   b. Physical exam
   c. Imaging tests
   a. Other_____________________
6. How do you assess the **degree of pain** in patients? **(**multiple-choice)
   a. Visual Analog Scale (VAS, scale 1-10)
   b. Numerical Pain Scale (NRS)
   c. According to the testimony of the patients (mild, moderate, severe).
   d. Other_____________________
7. What is the **pain experienced** by patients for whom you recommend NAC/LA/Br/Zn? (Single-choice)
   a. Mild pain, VAS <4
   b. Moderate pain, VAS 4-7
   c. Severe pain, VAS >7
   d. I recommend it for any of the above cases
8. Depending on the pain that patients experience, do you **adapt the dosage** of NAC/LA/Br/Zn? (Single-choice)
   a. Yes
   b. No
9. At the beginning of treatment with NAC/LA/Br/Zn, which concentration do you usually recommend as treatment for **primary dysmenorrhea**? (Single-choice)
   a. NAC 300 mg/day, LA 100 mg/day, Br 12.5 mg/day Zn 5 mg/day
   b. NAC 600 mg/day, LA 200 mg/day, Br 25 mg/day Zn 10 mg/day
   c. NAC 300 mg/day, LA 100 mg/day, Br 12.5 mg/day Zn 5 mg/day and, in case the patient doesn't respond, I double the concentrations per day.
   d. Other_____________________
10. When a patient is treated first with NAC/LA/Br/Zn, what is your **recommendation at the beginning of treatment for primary dysmenorrhea**? (Single-choice)
    a. 3 months of continuous treatment and re-evaluation.
    b. I recommend a long-term regimen, longer than 3 months.
    c. I recommend a short-duration regimen, less than 3 months.
    d. Other_____________________
11. If you initially recommend NAC/LA/Br/Zn with a **long-term regimen**, what regimen do you recommend for primary dysmenorrhea? (Single-choice)
    a. >3 months with 10-day breaks.
    b. >3 months without breaks.
    c. I recommend shorter time therapy at the beginning.
    d. Other_____________________
12. If you initially recommend NAC/LA/Br/Zn with a **short-duration regimen**, what regimen do you recommend for primary dysmenorrhea? (Single-choice)
    a. 5 days prior to menstruation.
    b. 7-15 days prior to menstruation.
    c. 5 days prior to menstruation and 5 days after menstruation.
    d. I do not usually recommend short-term treatment at the beginning.
    e. Other_____________________
13. Of the following options, which dosage do you recommend for patients with **primary dysmenorrhea with mild pain (<4)**? (Single-choice)
    a. 3 months continuous therapy with 10-day breaks.
    b. 3 months continuous therapy and change to 5 days before menstruation, (discontinuous therapy).
    c. 3 months continuous therapy and change to 7-15 days before menstruation (discontinuous therapy).
    d. 5 days before and 5 days after menstruation (discontinuous therapy).
    e. 7-15 days before menstruation (discontinuous therapy).
    f. Other_____________________
14. Of the following options, which dosage do you recommend for patients with **primary dysmenorrhea with moderate pain (4-7)**? (Single-choice)
    a. 3 months continuous therapy with 10-day breaks.
    b. 3 months continuous therapy and change to 5 days before menstruation, (discontinuous therapy).
    c. 3 months continuous therapy and change to 7-15 days before menstruation (discontinuous therapy).
    d. 5 days before and 5 days after menstruation (discontinuous therapy).
    e. 7-15 days before menstruation (discontinuous therapy).
    f. Other_____________________
15. Of the following options, which dosage do you recommend for patients **with primary dysmenorrhea with severe pain (>7)**? (Single-choice)
    a. 3 months continuous therapy with 10-day breaks.
    b. 3 months continuous therapy and change to 5 days before menstruation, (discontinuous therapy).
    c. 3 months continuous therapy and change to 7-15 days before menstruation (discontinuous therapy).
    d. 5 days before and 5 days after menstruation (discontinuous therapy).
    e. 7-15 days before menstruation (discontinuous therapy).
    f. Other_____________________
16. In case you **adapt the dosage because the response to treatment**, what do you recommend? (Single-choice)
    a. From continuous to discontinuous treatment, with treatment taken in the 5 days before menstruation.
    b. From continuous to discontinuous treatment, with treatment taken in the 7-15 days before menstruation.
    c. From continuous to discontinuous treatment, with treatment taken in the 5 days before and after menstruation.
    d. From discontinuous to continuous treatment
    e. Other_____________________
17. What do you think should be the **total duration of treatment** with NAC/LA/Br/Zn for patients with primary dysmenorrhea? (Single-choice)
    a. Indefinite
    b. Between 3 and 6 months
    c. <3 months
    d. Other_____________________
18. What do you consider to be the time needed to **evaluate the effectiveness** of NAC/LA/Br/Zn? (Single-choice)
    a. 15 days
    b. 30 days
    c. 90 days
    d. >90 days
    e. Other_____________________
19. Do you recommend NAC/LA/Br/Zn NAC/LA/Br/Zn in combination with **hygienic-dietary measures** for patients with dysmenorrhea? (Single-choice)
    a. No, only NAC/LA/Br/Zn
    b. NAC/LA/Br/Zn + regular physical exercise
    c. NAC/LA/Br/Zn + healthy diet
    d. NAC/LA/Br/Zn + avoid stress
    e. NAC/LA/Br/Zn + all measures mentioned above
20. If you recommend hygienic-dietary measures, do you **adapt the recommended NAC/LA/Br/Zn regimen**? (Single-choice)
    a. I do not change the dosage
    b. Yes, I change the dosage. Please indicate which one: _____________________

**BLOCK 3: SECONDARY DYSMENORRHEA AND OTHER CAUSES OF PAIN**

21. At the beginning of treatment with NAC/LA/Br/Zn, which concentration do you usually recommend as treatment for secondary dysmenorrhea associated with endometriosis? (Single-choice)
    a. NAC 300 mg/day, LA 100 mg/day, Br 12.5 mg/day Zn 5 mg/day
    b. NAC 600 mg/day, LA 200 mg/day, Br 25 mg/day Zn 10 mg/day
    c. NAC 300 mg/day, LA 100 mg/day, Br 12.5 mg/day Zn 5 mg/day and in case the patient doesn't respond, I double the concentrations per day.
    d. Other_____________________
22. When a patient is treated first with NAC/LA/Br/Zn, what is the **recommended duration of treatment** for secondary dysmenorrhea associated with endometriosis? (Single-choice)
    a. <3 months
    b. 3 months
    c. >3 months with 10-day breaks.
    d. >3 months without breaks.
    e. Other_____________________
23. For patients with endometriosis-associated secondary dysmenorrhea with **mild pain (<4)**, which dosage do you recommend? (Single-choice)
    a. 3 months continuous therapy with 10-day breaks.
    b. 3 months continuous therapy and change to 5 days before menstruation, (discontinuous therapy).
    c. 3 months continuous therapy and change to 7-15 days before menstruation (discontinuous therapy).
    d. 5 days before and 5 days after menstruation (discontinuous therapy).
    e. 7-15 days before menstruation (discontinuous therapy).
    f. Other_____________________
24. For patients with endometriosis-associated secondary dysmenorrhea with **moderate pain (4-7)**, which dosage do you recommend? (Single-choice)
    a. 3 months continuous therapy with 10-day breaks.
    b. 3 months continuous therapy and change to 5 days before menstruation, (discontinuous therapy).
    c. 3 months continuous therapy and change to 7-15 days before menstruation (discontinuous therapy).
    d. 5 days before and 5 days after menstruation (discontinuous therapy).
    e. 7-15 days before menstruation (discontinuous therapy).
    f. Other_____________________
25. For patients with endometriosis-associated secondary dysmenorrhea with **severe pain (>7)**, which dosage do you recommend? (Single-choice)
    a. 3 months continuous therapy with 10-day breaks.
    b. 3 months continuous therapy and change to 5 days before menstruation, (discontinuous therapy).
    c. 3 months continuous therapy and change to 7-15 days before menstruation (discontinuous therapy).
    d. 5 days before and 5 days after menstruation (discontinuous therapy).
    e. 7-15 days before menstruation (discontinuous therapy).
    f. Other_____________________
26. When the patient presents **dysmenorrhea associated with other pathologies** (e.g., chronic pelvic pain, myoma), do you recommend NAC/LA/Br/Zn as a treatment? (Single-choice)
    a. Yes
    b. No, only for endometriosis and/or dysmenorrhea associated with endometriosis
    c. Other________________
27. If you recommend NAC/LA/Br/Zn to a patient with **dysmenorrhea associated with other pathologies**, what dosage do you usually recommend? (Single-choice and NOT mandatory)
    a. 3 months continuous therapy with 10-day breaks.
    b. 3 months continuous therapy and change to 5 days before menstruation, (discontinuous therapy).
    c. 3 months continuous therapy and change to 7-15 days before menstruation (discontinuous therapy).
    d. 5 days before and 5 days after menstruation (discontinuous therapy).
    e. 7-15 days before menstruation (discontinuous therapy).
    f. Other_____________________

**BLOCK 4: ASSOCIATIONS WITH OTHER DRUGS**

28. In patients treated with **non-steroidal anti-inflammatory drugs (NSAIDs)**, what dosage of NAC/LA/Br/Zn do you recommend? (Single-choice)
    a. I do not recommend NAC/LA/Br/Zn
    b. I do not modify the dosage
    c. I recommend reducing the concentration of NAC/LA/Br/Zn
    d. Other dosage. Please indicate the dosage: _____________________
29. In patients treated with **combined hormonal contraception**, what dosage of NAC/LA/Br/Zn do you recommend? (Single-choice)
    a. I do not recommend NAC/LA/Br/Zn
    b. I do not modify the dosage
    c. I recommend reducing the concentration of NAC/LA/Br/Zn
    d. Other dosage. Please indicate the dosage: _____________________
30. In patients treated with **gestagens** (norethisterone, Dienogest, IUD), what dosage of NAC/LA/Br/Zn do you recommend? (Single-choice)
    a. I do not recommend NAC/LA/Br/Zn
    b. I do not modify the dosage
    c. I recommend reducing the concentration of NAC/LA/Br/Zn
    d. Other dosage. Please indicate the dosage: _____________________
31. In patients treated with **hypoglycemic drugs**, what dosage of NAC/LA/Br/Zn do you recommend? (Single-choice)
    a. I do not recommend NAC/LA/Br/Zn
    b. I do not modify the dosage
    c. I recommend reducing the concentration of NAC/LA/Br/Zn
    d. Other dosage. Please indicate the dosage: _____________________

**BLOCK 5: PATIENT SATISFACTION**

32. How often do you usually do a **follow up of patients** **with dysmenorrhea** to whom you have recommended NAC/LA/Br/Zn? (Single-choice)
    a. Once or twice a month
    b. Once every three months
    c. Once every six months
    d. Other_____________________
33. According to the feedback of your patients treated with NAC/LA/Br/Zn, do you **adapt or modify the dosage**? (Single-choice)
    a. Yes
    b. No
    c. Other reasons _____________________
34. If you adapt the treatment regimen, **what dosage do you usually change to**? (Single-choice)
    a. To continuous treatments (>3 months).
    b. To discontinuous treatments 5 days prior to menstruation.
    c. To discontinuous treatments 7-15 days prior to menstruation.
    d. To discontinuous treatments 5 days before and after menstruation.
    e. Other_____________________
35. Have any patients reported **ineffectiveness** **after the first 15 days** **of treatment** with NAC/LA/Br/Zn (Single-choice)?
    a. Yes
    b. No
36. Has any patient reported any **adverse reaction** during treatment with NAC/LA/Br/Zn? (Single-choice)
    a. Yes
    b. No
37. If any patient reported any **adverse reaction**, could you please indicate which one? (Short answer):

____________________________________________________

38. If you have any suggestions or comments, please feel free to express them in the box below.

Thank you very much for your contribution and for your participation.
