## Supplementary Figure for "Study of the recommended dosage of the N-Acetyl Cysteine, Alpha Lipoic Acid, Bromelain and Zinc preparation as a treatment for dysmenorrhea"

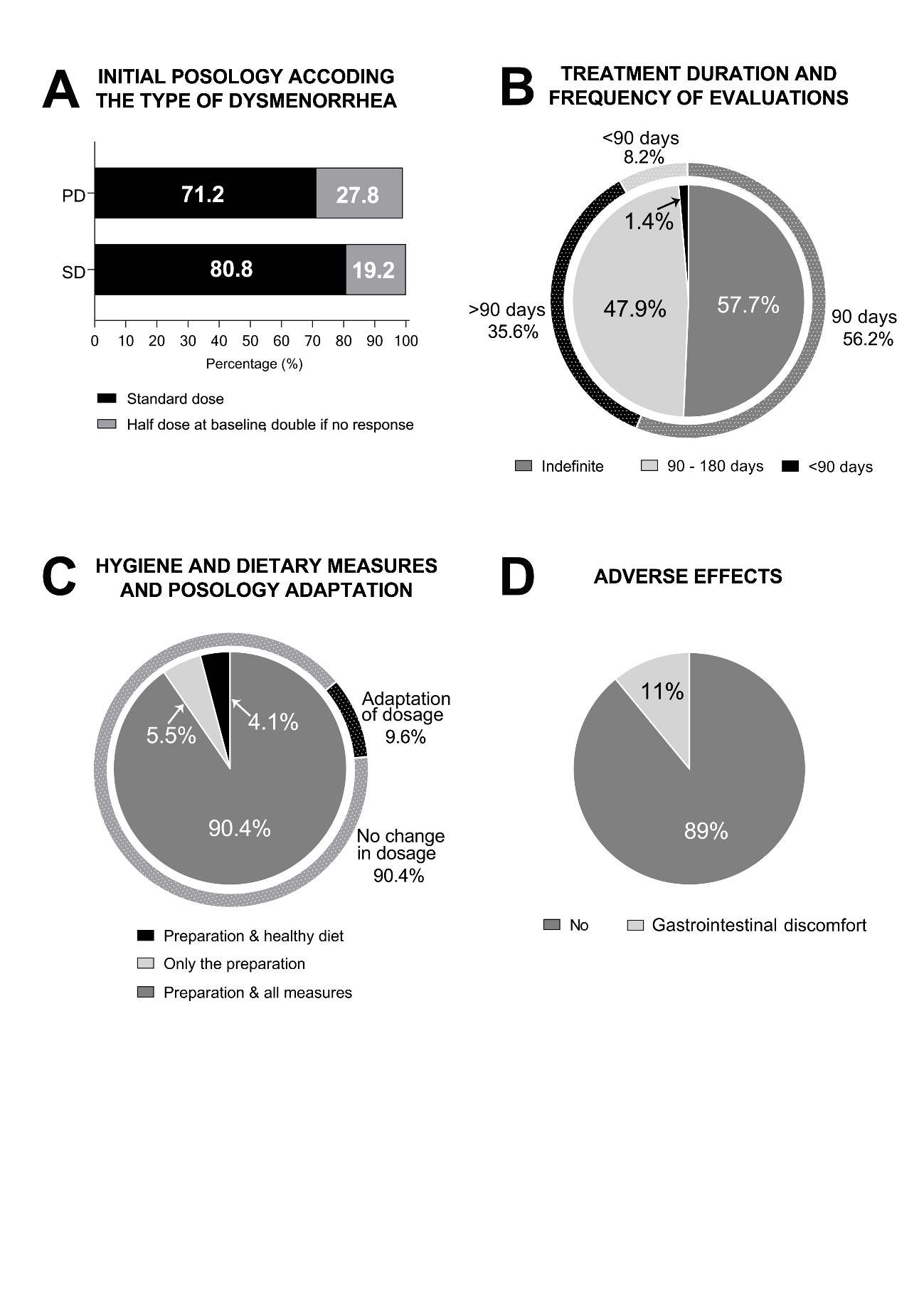

**Supplementary Figure: (A)** Initial dosage for primary dysmenorrhea (PD) and secondary dysmenorrhea (SD), considering NAC 600 mg/day; LA, 200 mg/day; Br 25 mg/day; Zn, 10 mg/day as standard dose. Initial posology recommendation for PD and SD. **(B, inner chart)** Duration of NAC/LA/Br/Zn treatment; **(B, outer ring)** frequency of evaluations. **(C, inner chart)** Hygiene and dietary measures recommendations and its association with the preparation (Preparation and all measures (healthy diet, regular exercise and avoiding stress) 90.4%; only the preparation 5.4%; Preparation and healthy diet 4.1%.); **(C, outer ring)** Expert adaptation of the preparation posology regarding the hygiene and dietary measures recommendations (No change in dosage 90,4%; Adaptation of dosage 9.6%). **(D, inner chart**) Adverse effects reported by the patients (No 89%; 11% Yes). (**D, outer ring**) Adverse effect reported by specialist (89% no adverse effect; 10.9% gastrointestinal discomfort). All results are represented as percentage, N=73.
